# Thalamic volume as a susceptibility/risk biomarker for *C9orf72* phenoconversion

**DOI:** 10.1101/2025.09.20.25336234

**Authors:** Ting Shen, Joanne Wuu, Christopher A. Olm, Hamsanandini Radhakrishnan, Yindi Li, Pradip M. Pattany, Volkan Granit, Nathan Carberry, David J. Irwin, Lauren Massimo, Defne A. Amado, Colin Quinn, Michael Baer, Laynie Dratch, Lauren Elman, Michael Benatar, Corey T. McMillan

## Abstract

**Background:** *C9orf72* repeat expansions are the most frequent genetic risk factor of amyotrophic lateral sclerosis (ALS) and frontotemporal degeneration (FTD). With growing interest in the prospect of preventing clinically manifest disease, there is a pressing need to identify susceptibility/risk biomarkers that might predict the short-term risk of phenoconversion. This study sought to identify such neuroimaging biomarkers that predict the risk of ALS or FTD among unaffected *C9orf72* carriers.

**Methods:** Our cohort comprised 4 groups of participants genotyped for *C9orf72* repeat expansions, including 73 pre-symptomatic carriers, 8 phenoconverters, 49 affected carriers, and 99 non-carrier controls. Pre-symptomatic carriers remained unaffected throughout the follow-up period, while phenoconverters were those who transitioned from pre-symtpomatic to the clinically manifest phase during follow-up. Affected carriers were already diagnosed with ALS and/or FTD at baseline. All participants underwent an initial MRI scan, with a subset receiving follow-up MRI scans. Regional gray matter volumes were analyzed to assess initial differences across groups, to predict phenoconversion from a pre-symptomatic to clinically manifest state, and to explore progression over time.

**Results:** Group comparisons revealed an increasing extent and magnitude of reduced gray matter volume across the clinical continuum, from pre-symptomatic to clinically manifest stages, with phenoconverters who were imaged when pre-symptomatic and later developed clinically manifest disease intermediate between the two. The thalamus demonstrates the largest effect and the least variability across centers and MRI protocols. Thalamic volume is, on average, lower among pre-symptomatic carriers than non-carrier controls, though with wide overlapping distributions. Phenoconverters exhibited thalamic volumes in the range of affected carriers, both before and after phenoconversion. Mean thalamic volume discriminated between phenoconverters and carriers who remained pre-symptomatic during follow-up with an area under the curve of 0.854 (p<0.001), and time-to-phenoconversion analysis demonstrated that individuals with a lower baseline thalamic volume had an increased hazard for phenoconversion (HR=17.6; CI=2.2-143.3; p<0.001). Other regions, including the amygdala, somatomotor cortex, postcentral gyrus, and parietal cortex, demonstrated less consistent signals across centers and MRI protocols, but generally followed trends similar to the thalamus. Longitudinal observations further indicated that these regions, particularly the thalamus, demonstrated consistent downward trajectories over time, with more rapid atrophy observed in phenoconverters and affected carriers.

**Conclusions:** Lower thalamic volume is a promising susceptibility/risk biomarker predicting phenoconversion to clinically manifest ALS or FTD among clinically unaffected *C9orf72* repeat expansion carriers, with potential utility to aid the design and implementation of early intervention and preventative clinical trials.

## Introduction

A heterozygous CCCCGG hexanucleotide expansion in the non-coding region of *C9orf72* gene is the major genetic cause of autosomal dominant amyotrophic lateral sclerosis (ALS) and frontotemporal degeneration (FTD)^1^, irrespective of family history^2^. As we enter an era of prevention trials in genetic ALS and FTD, clinically unaffected individuals with a *C9orf72* expansion represent a well-defined population in which early therapeutic intervention might be possible^3^. A major barrier to *C9orf72* prevention trials, however, is the current limited ability to predict when clinically manifest disease (ALS and/or FTD) will develop among those with a *C9orf72* repeat expansion. Thus, there is a critical need to identify *susceptibility/risk* biomarkers that are measurable in pre-symptomatic at-risk individuals and that predict the future risk of developing clinically manifest disease.

There is substantial evidence that MRI is sensitive to the detection of structural neurodegeneration prior to clinically manifest ALS-FTD^4,5^. However, the vast majority of studies report group-level results and lack a clear focus on how structural MRI changes might be utilized as a biomarker with a defined context of use. Despite these limitations, smaller thalamic volume has emerged as a potential early biomarker of pre-symptomatic disease among those with a *C9orf72* repeat expansion. An early cross-sectional study, for example, suggested that lower thalamic volume might precede the emergence of clinically manifest disease by ∼20 years^5^, but this analysis was predicated on the idea that age of clinical symptom onset could reliably be estimated based on age of onset among family members, which is now known to be unreliable^6^. Subsequent, relatively small, cross-sectional studies have demonstrated lower thalamic volume among unaffected individuals with a *C9orf72* repeat expansion^7-9^. However,.despite cross-sectional evidence of thalamic volume loss as many as 6 years before the emergence of clinically manifest disease, longitudinal analyses indicate that the rate of thalamic volume change in pre-symptomatic carriers does not significantly differ from non-carriers^4^

From the perspective of a trial that aims to prevent clinically manifest disease among *C9orf72* carriers, a key question is whether thalamic volume among clinically unaffected carriers with a C9orf72 expansion might have value as a susceptibility/risk biomarker, predicting the time to phenoconversion to clinically manifest disease, irrespective of whether the initial emergent phenotype is ALS or FTD.

This was the focus of this current multi-center study specifically investigating the hypothesized utility of the thalamus, but also exploring additional neuroanatomical regions.

## Methods

### Study Oversight

This study involving human participants was reviewed and approved by the institutional review boards of the University of Pennsylvania (IRB protocol #842873) and University of Miami (IRB protocol #20101021). Written informed consent was obtained from all the participants or caregivers in accordance with the Declaration of Helsinki. Ethical approval was given and is effective.

### Study Cohorts

This study includes research participants enrolled in either the University of Pennsylvania Centralized Observational Research Repository on Neurodegenerative Disease (UNICORN) study or the University of Miami Pre-Symptomatic Familial ALS (*Pre-fALS*) and Clinical Research in ALS (CRiALS) Biomarker studies^10^. The UNICORN Study is a 20-year longitudinal natural history study that follows unaffected individuals at genetic risk for FTD or ALS who are in families with known pathogenic variants in *C9orf72, GRN, MAPT*, or other less common genes (e.g., *VCP*). Participation involves collection of DNA sample, plasma, MRI, and clinical evaluation on a semi-annual basis. Participants are offered optional genetic counseling and predictive testing, but genetic status is not required to be known for participation. As such, we follow individuals longitudinally with and without confirmed genetic risk or participants’ knowledge of their genetic status. In addition to research participants who are unaffected at baseline, the UNICORN study includes individuals with symptomatic FTD and/or ALS, and we continue to longitudinally characterize individuals who undergo phenoconversion from pre-symptomatic to clinically manifest disease under the same protocol. *Pre-fALS*, initiated in 2007, is a natural history and biomarker study that recruits individuals at genetic risk of ALS or ALS-FTD (i.e., carriers of pathogenic variants in *SOD1, C9orf72, FUS, TARDBP, VCP, etc*.) from across North America. Participants undergo genetic counseling and testing prior to enrollment, with the opportunity to learn their genetic status (disclosure group) or not (non-disclosure group), if genetic status is not already known (known status group). The non-disclosure groups include both individuals with and without the pathogenic variant(s) responsible for disease in their family. All participants are unaffected at the time of enrolment and undergo MRI during the pre-symptomatic stage of disease, around the time of phenoconversion to either ALS or FTD, and early into the clinically manifest stage of disease. The CRiALS Biomarker study recruits healthy controls and patients with ALS; study participants undergo the same longitudinal study procedures as in *Pre-fALS*.

For this analysis, we included participants who had been genotyped for a *C9orf72* repeat expansion with a threshold for defining a pathologically expanded repeat length determined based on local laboratory standards, each of whom was designated as belonging to one of four groups: (a) unaffected family members who do not have a *C9orf72* repeat expansion (controls); (b) carriers of a *C9orf72* repeat expansion who remained clinically unaffected, based on UNICORN or *Pre-fALS* assessments, throughout follow-up (pre-symptomatic carriers); (c) those who were followed from the pre-symptomatic stage through phenoconversion to the clinically manifest stage of disease (phenoconverters); and (d) those who were already affected with either ALS or FTD at the time of enrollment (clinically manifest). For inclusion in this analysis, participants were required to have undergone a T1-weighted MRI.

### Genetic screening

Genomic DNA was extracted from peripheral blood or brain tissue collected from participants^11^. Genotyping for *C9orf72* hexanucleotide repeat expansions was performed using a modified repeat-primed polymerase-chain reaction, as previously described ^12^. We defined a pathogenic *C9orf72* repeat expansion as >30 repeats^13^.

### Neuroimaging acquisition

Structural T1-weighted (T1w) MRI scans were acquired on Siemens 3.0 Tesla scanners. Given the follow-up duration of up to ∼20 years, the T1-weighted MRI scans were collected with several similar magnetization-prepared rapid gradient-echo (MPRAGE) sequences as follows: (1) TIM Trio scanner, 8 channel head coil, axial plane with repetition time (TR) = 1620 ms, echo time (TE) = 3.09 ms, slice thickness = 1.0 mm, in-plane resolution = 0.98 × 0.98 mm. (2) TIM Trio scanner, 64 channel head coil, sagittal plane with TR = 2300 ms, TE = 2.95 ms, slice thickness = 1.2 mm, in-plane resolution = 1.05 × 1.05 mm. (3) TIM Trio scanner, 64 channel head coil, sagittal plane with TR = 2300 ms, TE = 2.98 ms, slice thickness = 1.0 mm, in plane resolution = 1.0 × 1.0 mm. (4) Prisma scanner, 64 channel head coil, sagittal plane with TR = 2400 ms, TE = 1.96 ms, slice thickness = 0.8 mm, in-plane resolution = 0.8 × 0.8 mm. (5) TIM Trio scanner, 8 channel head coil, axial plane with TR = 2300 ms, TE = 2.4 ms, slice thickness = 1.0 mm, in plane resolution = 1.0 × 1.0 mm. We covary for each MRI sequence in all analyses.

### Neuroimaging Processing

We first prepared the T1-weighted MRI images for processing by reorienting all images to left, posterior, and inferior (LPI) orientation and performed neck-trimming to reduce the number of “extra” slices in the images to minimize bias in field-of-view across the different acquisition sequences. The neck trimming also served to decrease image size, which helps to optimize the performance of our computing resources. The T1-weighted images were then processed using SynthSeg (https://github.com/BBillot/SynthSeg)^14^. SynthSeg was trained and validated on data acquired with various image types, resolutions, and contrasts from an assortment of scanners with the stated purpose of applying it to heterogenous datasets, making it well-suited to use for this multi-site study. Briefly, SynthSeg resamples images to 1 mm^3^ isotropic resolution and then uses a neural network to segment images, and in particular the subcortical gray matter structures and cortical regions from the freesurfer atlas^15^. We extracted cubic mm volumes from these structures and divided them by the total intracranial volume (ICV) before analysis.

### Statistical analysis

Statistical analyses and plotting were carried out using R statistical software (version 4.1.0). The brain heatmaps were visualized using BrainNet Viewer^16^. There were four groups in this study, including non-carriers, pre-symptomatic carriers, phenoconverters, and affected carriers. To investigate gray matter structure in baseline MRIs across groups, a linear model was fitted with group as a fixed effect and covariates including age at the time of initial MRI, sex and MRI sequence. This model was used to evaluate regional differences in ICV-corrected gray matter volume at initial MRI scan, comparing pre-symptomatic carriers, phenoconverters, and affected carriers to non-carriers of the *C9orf72* expansion. Separately for each of the four groups, correlations of regional ICV-corrected gray matter volume with age were assessed using a mixed effects model that were adjusted for sex and MRI sequence, and accounting for within-person correlations. False discovery rate (FDR) correction was applied to control for multiple comparisons, and a significance level of FDR-corrected p < 0.05 was considered as statistically significant. To visualize relationships between gray matter volume and age while accounting for the covariates, partial regression plots were generated, with the residuals of gray matter volume (after regressing out sex and MRI sequence, and accounting for within-person correlations) plotted against the residuals of age (likewise adjusted for sex and MRI sequence). Building on these analyses, we further added the age*group interaction term to the linear model to test significant age x group interaction effects in regions that showed substantial volume alterations in carriers of a *C9orf72* expansion.

To evaluate the predictive utility of regional gray matter volume as a susceptibility/risk biomarker of phenoconversion in indivudials with a *C9orf72* expansion, we first performed receiver operating characteristic (ROC) analysis in pre-symptomatic carriers (those who remain unaffected throughout the entire study period) and phenoconverters (those who convert to manifest ALS and/or FTD during the study period) using ICV-corrected gray matter volume derived from the initial available MRI of each participant as the predictor. The optimal volume threshold for distinguishing subsequent phenoconverters was determined by maximizing Youden’s index, and its sensitivity and specificity were calculated. Participants were then stratified into high-risk (volume below threshold) and low-risk (volume above threshold) groups. Then we conducted Kaplan-Meier survival analysis with log-rank testing to compare phenoconversion probability between risk groups.

## Results

### Participant characteristics

We included a total of 229 participants – 99 controls, 73 pre-symptomatic carriers, 8 phenoconverter carriers, and 49 clinically-manifest carriers who had ALS and/or FTD at their baseline scan.

Demographic and clinical characteristics of each group are summarized in Table 1. There were no significant differences in age, sex or education between pre-symptomatic carriers and non-carrier controls. Phenoconverters and affected carriers were older and had a higher proportion of males compared to the other two groups. Among the 8 phenoconverters, 3 developed motor symptoms and progressed to ALS, and 5 initially presented with cognitive and behavioral symptoms and then progressed to FTD during the follow-up period. Of the 49 affected carriers in our cohort at the time of initial MRI, 31 had FTD, 10 had ALS and 8 had ALS-FTD. A subset of 113 participants had at least one additional scan, with all follow-up intervals exceeding a minimum of 6 months.

**Table 1.** Demographic and clinical characteristics of the study population.

|  | **Non-carriers** | **Pre-symptomatic** | **Phenoconverters** | **Clinically Manifest** |
| --- | --- | --- | --- | --- |
| **Number of participants, N** | 99 | 73 | 8 | 49 |
| **Contributing site, N (%)** |  |  |  |  |
| Penn | 67 (67.7%) | 23 (31.5%) | 2 (25.0%) | 48 (98.0%) |
| Miami | 32 (32.3%) | 50 (68.5%) | 6 (75.0%) | 1 (2.0%) |
| **Sex, N (%)** |  |  |  |  |
| Male | 38 (38.4%) | 26 (35.6%) | 5 (62.5%) | 29 (59.2%) |
| Female | 61 (61.6%) | 47 (64.4%) | 3 (37.5%) | 20 (40.8%) |
| **Education, Mean (SD) years** | 15.9 (2.4) | 16.1 (2.3) | 14.9 (2.8) | 15.6 (3.0) |
| **Age at initial scan, Mean (SD) years** | 42.4 (12.8) | 43.1 (11.8) | 58.5 (7.4) | 60.9 (8.0) |
| **Age at symptom onset, Mean (SD) years** | (n/a) | (n/a) | 62.7 (8.7) | 57.2 (7.7) |
| **Time from symptom onset to initial MRI, Mean (SD) years** | (n/a) | (n/a) | -4.6 (3.5) | 3.6 (3.1) |
| **Clinical phenotype,<sup>1</sup> N (%)** |  |  |  |  |
| ALS | (n/a) | (n/a) | 3 (37.5%) | 10 (20.4%) |
| ALS-FTD | (n/a) | (n/a) | 0 (0.0%) | 8 (16.3%) |
| FTD | (n/a) | (n/a) | 5 (62.5%) | 31 (63.2%) |
| **Onset symptom, N (%)** |  |  |  |  |
| Motor | (n/a) | (n/a) | 3 (37.5%) | 17 (34.7%) |
| Cognitive/Behavioral | (n/a) | (n/a) | 5 (62.5%) | 32 (65.3%) |
| **Longitudinal MRI subset** |  |  |  |  |
| **Number of participants, N (%)** | 37 (37%) | 47 (64%) | 6 (75%) | 23 (47%) |
| **Total number of MRI scans, Median (Range)** | 3 (2-5) | 2 (2-6) | 4.5 (2-6) | 3 (2-7) |
| **Follow-up period (from initial scan to the last follow-up scan), Median (Q1-Q3) years** | 4.2 (2.5-5.4) | 2.5 (1.6-4.8) | 4.7 (2.0-6.5) | 1.9 (1.1-3.5) |

<sup>1</sup> Clinical phenotype reflects the first observed clinically manifest syndrome for both the phenoconverter and clinically manifest groups.

### Gray matter volume comparisons

We compared first available, “baseline”, gray matter volumes across groups, adjusting for age, sex, and MRI sequence. As expected, relative to healthy controls without a *C9orf72* expansion, pre-symptomatic carrier, phenoconverter, and affected carrier groups each showed evidence of reduced thalamic volume. In the affected group, in addition to bilateral thalamic volume, there was evidence for reduced volume in hippocampus and amygdala limbic regions and widespread neocortex, particularly in frontal regions (Fig.1C). Pre-symptomatic carriers displayed reduced volume in bilateral thalamus, right amygdala, as well as several cortical regions, including the left inferior and superior parietal cortex, bilateral middle temporal cortex, left pars triangularis, bilateral precuneus, bilateral supramarginal gyrus, and right rostral middle frontal cortex (Fig.1A). The distribution of volumetric changes among pre-symptomatic carriers represents a subset of regions identified in the affected group, but with less pronounced reductions in volume.

**Figure 1.**
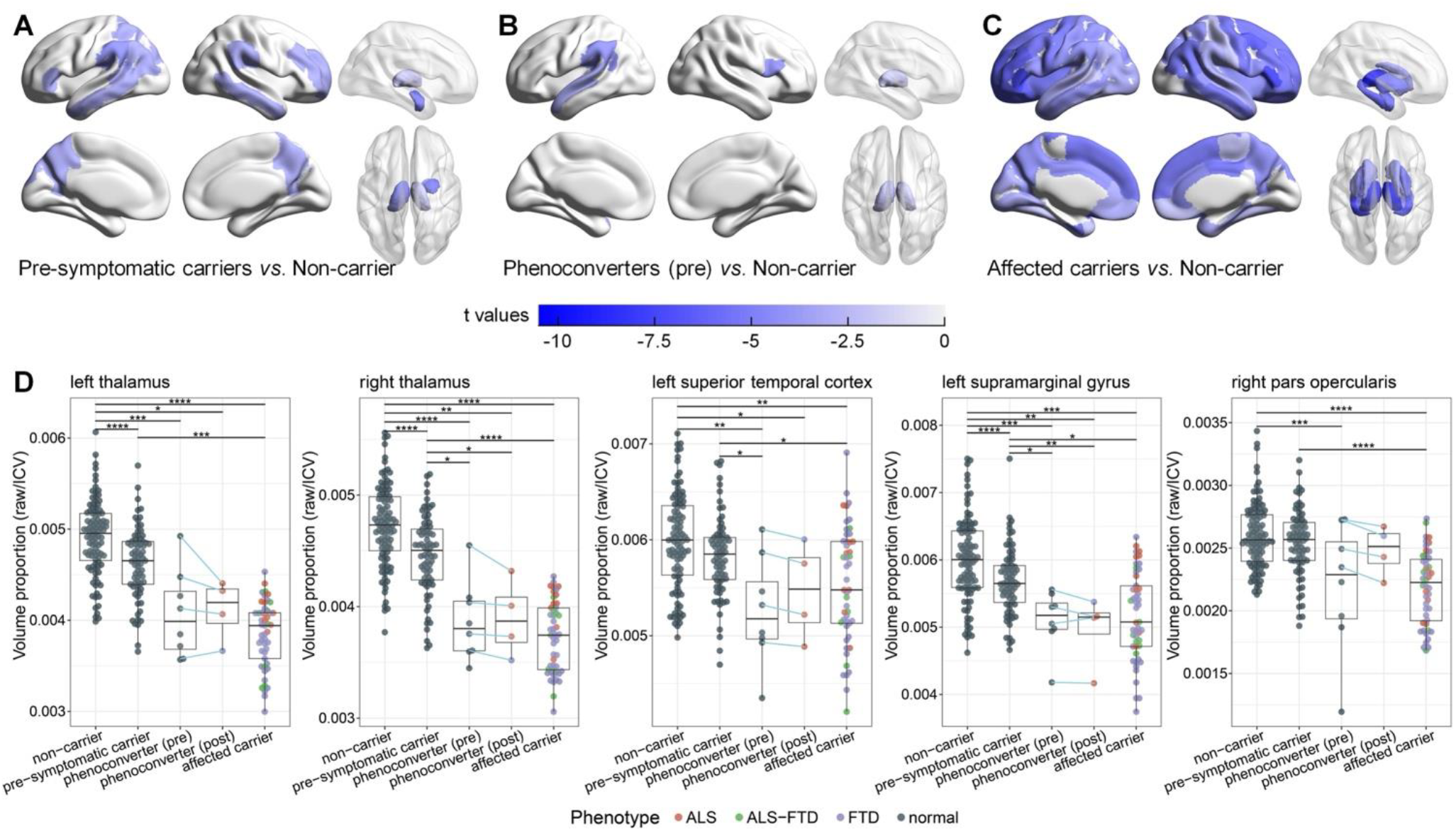
Gray matter volume in non-carrier controls and carriers of a *C9orf72* repeat expansion. Comparison of gray matter volume across groups: pre-symptomatic carriers (**A**), phenoconverters (**B**), and affected carriers (**C**) relative to non-carrier healthy controls. Only regions with FDR-corrected p < 0.05 are displayed. Darker blue shading represents more substantial volume reductions. (**D**) Volumetric values of significantly reduced regions across groups. * indicates p < 0.05, ** indicates p < 0.01, *** indicates p < 0.001, **** indicates p < 0.0001. Affected individual dots colored to reflect initial clinical diagnosis: ALS (red), FTD (purple), ALS-FTD (green).

Relative to non-carriers, lower volume on the initial pre-symptomatic scans among phenoconverters were apparent in the thalamus bilaterally as well as the left superior temporal cortex, left supramarginal gyrus, and right pars opercularis cortical regions (Fig. 1B). The distribution of individual-level differences in volumetric measures across participant groups (controls, pre-symptomatic, phenoconverters pre- and post-conversion, and clinically manifest) is further illustrated in Fig. 1D. The observed pattern is consistent with a progressive group-level reduction in thalamic volumes bilaterally across the phenotypic continuum, with the most pronounced reduction in the clinically manifest stage. Volumetric measures in the phenoconverters were intermediate between pre-symptomatic and clinically manifest groups, with individual-level longitudinal changes from 4 phenoconverters who underwent an MRI both before and after phenoconversion illustrated with connecting lines. While not evaluated statistically due to limited cases, the observed pattern is one of modest decline in each of the evaluated regions, most pronounced in the thalamus bilaterally. Moreover, while the lowest volumes were apparent in FTD followed by ALS-FTD, individual-level volumes are largely overlapping, suggesting these volumetric signatures of *C9orf72* are relatively independent of whether the clinical phenotype manifest in the motor neuron or frontotemporal axis^17^.

### Correlations of regional gray matter volume with age

While it is not yet possible to reliably estimate expected age of onset of clinical manifestations of disease among at-risk individuals with a *C9orf72* expansion, penetrance is age-dependent and increases with advancing age^18^. It was, therefore, necessary to investigate age-related effects. Across all groups we observed widespread gray matter volume reductions in association with advancing age (Fig. 2). As expected, age-related gray matter volume decline was especially prominent in frontal and temporal cortical regions, as well as subcortical structures, suggesting selective vulnerability of these areas to the aging processes. This pattern was evident across the *C9orf72* phenotypic continuum. In healthy controls without expansions (age range = 19.2 - 74.6 years), we observed the strongest inverse correlations between increased age and lower gray matter volume in the thalamus bilaterally (Fig. 2A). Notably, this pronounced thalamic age-volume relationship persisted with comparable magnitudes in both pre-symptomatic carriers (Fig. 2B) and phenoconverters (Fig. 2C). Affected carriers (age range = 39.6 - 76.0 years) also demonstrated significant age-volume correlations across a wide range of brain regions including the thalamus, despite narrower chronological age spread (Fig. 2D).

**Figure 2.**
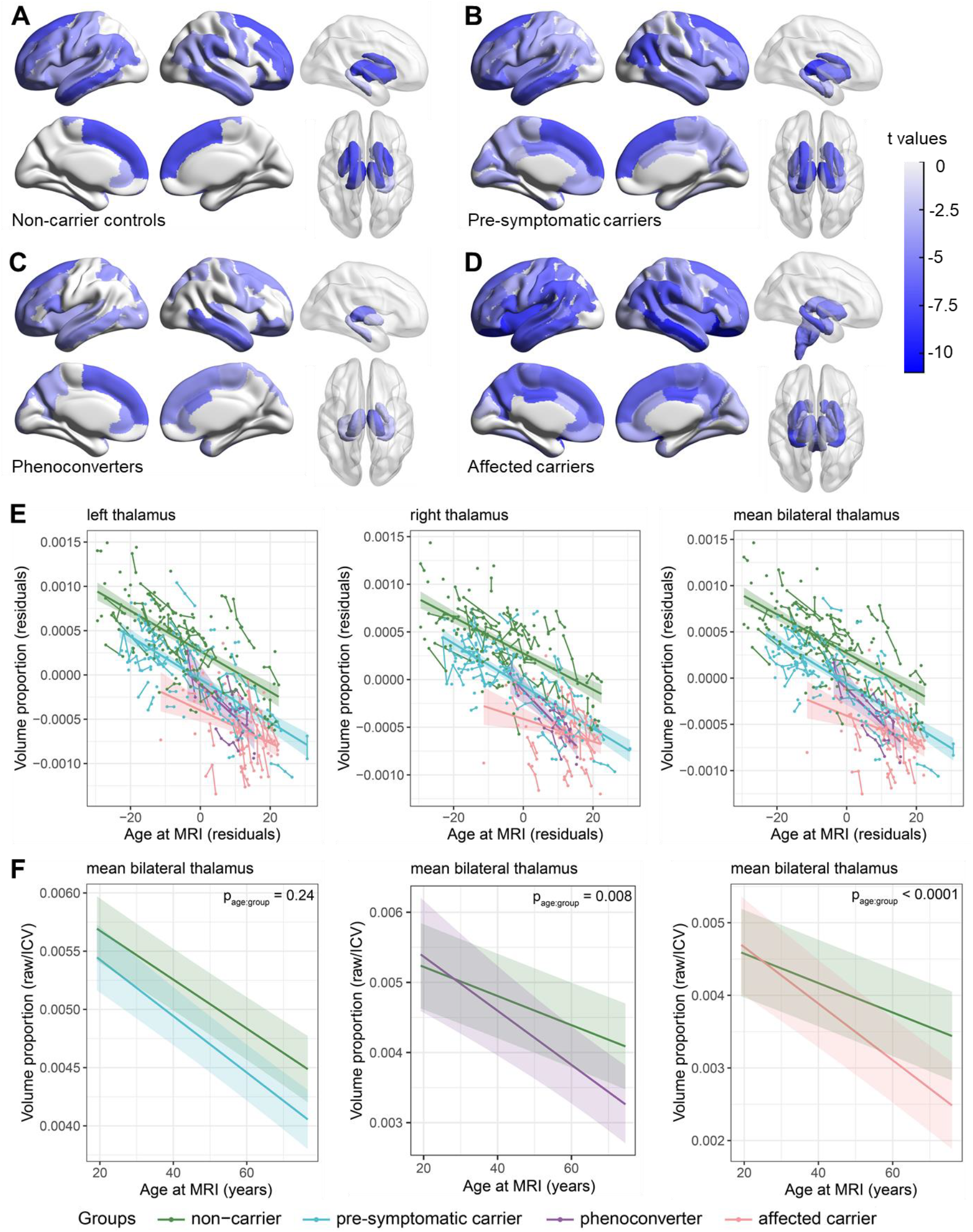
Correlations of regional gray matter volume with age across groups. (**A-D**) Heatmap showing correlations between gray matter volume and age across brain regions. Only regions with FDR-corrected p < 0.05 are displayed. Darker blue indicates stronger statistical significance. (**E**) Partial regression plots of thalamic volume versus age at MRI after adjusting for sex and MRI sequence. Points represent residual thalamic volume proportions (y-axis) and residual ages (x-axis) for each participant. Regression lines and 95% confidence intervals are shown for each group. (**F**) Age-related trajectories of mean thalamic volume by each groups for independent interaction analyses. Lines and shaded 95% confidence intervals from a linear mixed model accounting for sex and MRI sequence. P_age:group_ represents the significance of age-by-group interactions.

After adjusting for sex and MRI sequence effects and adjusting for within-person correlation, partial regression plots generated using data from all scans, revealed that both pre-symptomatic and affected carriers exhibited patterns of age-related thalamic volume decline similar to controls, though affected carriers exhibited more pronounced volume reduction (Fig. 2E). Notably, phenoconverters showed steeper decline slopes in the thalamus bilaterally, compared to other groups, indicating accelerated volumetric change beyond normative aging. Despite limited sample size, this directional pattern suggests disease-associated volumetric reductions in the thalamus compounded with age-related decline. Supporting this interpretation, phenoconverters’ trajectory originated within the confidence intervals of pre-symptomatic carriers and progressively descended to overlap with affected carriers’ confidence intervals.

To disentangle disease-related and age-related volumetric patterns, we examined age-by-group interaction effects on thalamic volumes in pre-symptomatic carriers, phenoconverters, and affected carriers relative to non-carriers, while adjusting for sex and MRI sequences and accounting for within-person correlations (Fig. 2F, Supplementary Fig. 1). Considering mean thalamic volume (averaged across hemispheres), we did not observe an interaction between age and group for pre-symptomatic carriers and non-carriers, who on average exhibited parallel trajectories of thalamic volumetric decline across age (Fig.2F). By contrast, an age x group interaction showed that phenoconverters (p=0.008) and affected carriers (p<0.0001) have significantly greater thalamic volume reductions across age compared to non-carriers. These patterns suggest that the reduction in thalamic volume in these two groups is not simply a reflection of age alone, but exaggerated as a function of underlying disease.

### Phenoconversion prediction using thalamic volume

To empirically determine whether thalamic volume can serve as a susceptibility/risk biomarker of phenoconversion among unaffected *C9orf7*2 carriers, we performed ROC analysis using the first available MRI scan from pre-symptomatic carriers and phenoconverters prior to phenoconversion (Fig. 3A). The mean thalamic volume, averaged across left and right sides and normalized to ICV, showed excellent discriminative value in identifying those *C9orf72* carriers who subsequently phenoconverted compared to those who did not (AUC = 0.854, p < 0.001). At an optimal thalamic volume proportion (thalamic volume/ICV) cutoff of 0.0043, the model achieved 83% sensitivity and 79% specificity. Using this threshold, we stratified participants into high-risk group (thalamic volume proportion < 0.0043) and low-risk group (thalamic volume proportion > 0.0043). Kaplan-Meier survival analysis showed significantly higher phenoconversion rates in the high-risk group compared to low-risk participants (HR=17.6, CI=2.2-143.3; log-rank p = 0.0002; Fig. 3B). This predictive pattern remained consistent when analyzing left and right thalamus separately (Supplementary Fig. 2), and when adjusting for age in the model (Supplementary Fig. 3), demonstrating the robustness of thalamic volume as a susceptibility/risk biomarker.

**Figure 3.**
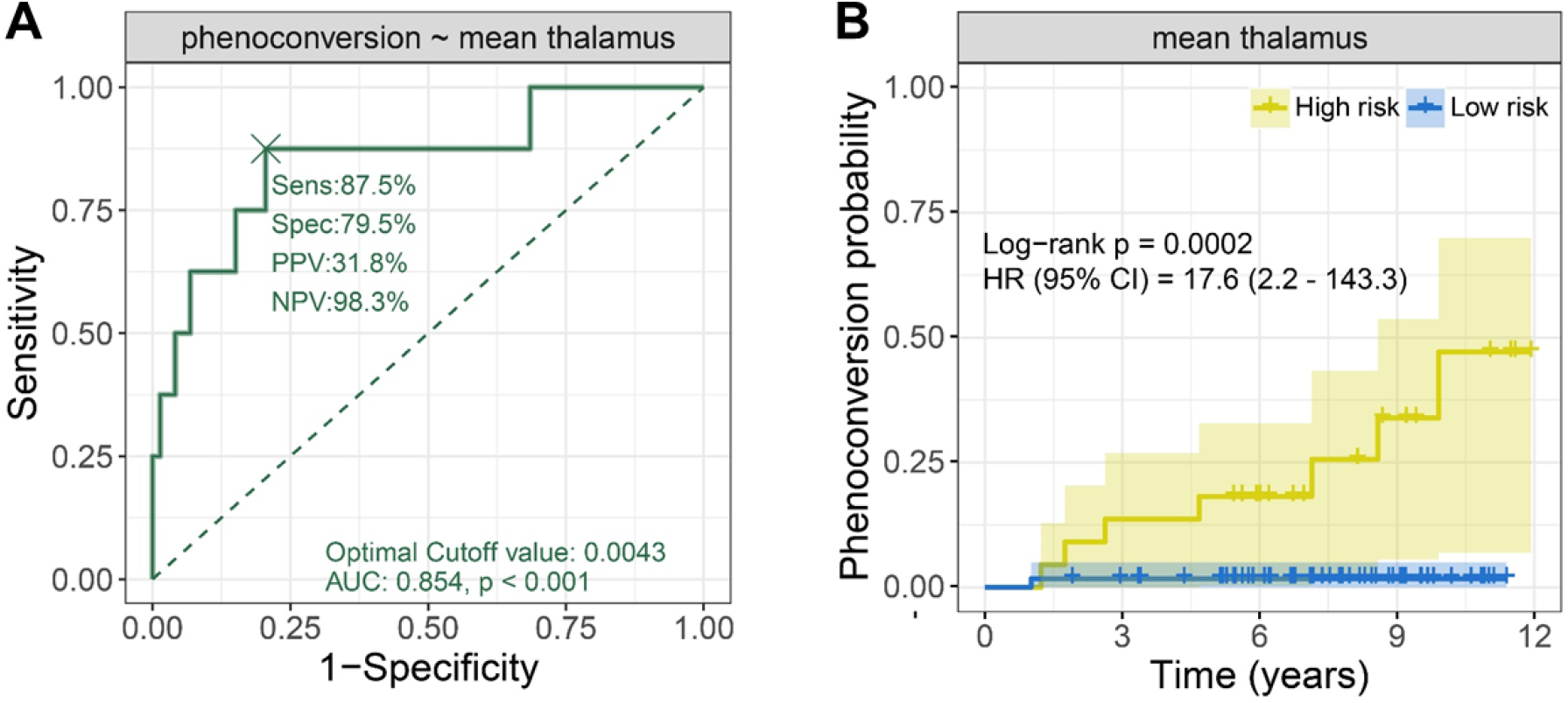
Analysis of phenoconversion prediction. (**A**). ROC curves for discriminating phenoconversion using mean bilateral thalamic volume. The “×” mark represents the optimal cutoff point. (**B**) Kaplan-Meier curves showing phenoconversion probability by risk groups. Shading represents 95% CI. HR, hazard ratio; CI, confidence interval.

### Longitudinal changes of gray matter volume

Our primary investigation of thalamic volume as a susceptibility/risk biomarker utilized cross-sectional (initial scan) MRI data. Secondarily, we evaluated longitudinal changes in the thalamus in the phenoconverters and affected-carriers (Fig. 4). The thalamus exhibited consistent downward trajectories as the disease progressed in both phenoconverters and affected carriers. When aligned to years to/from symptom onset, longitudinal trajectories revealed that volumetric decline begins prior to phenococonversion and continues on a downward trajectory over the course of clinically manifest disease.

**Figure 4.**
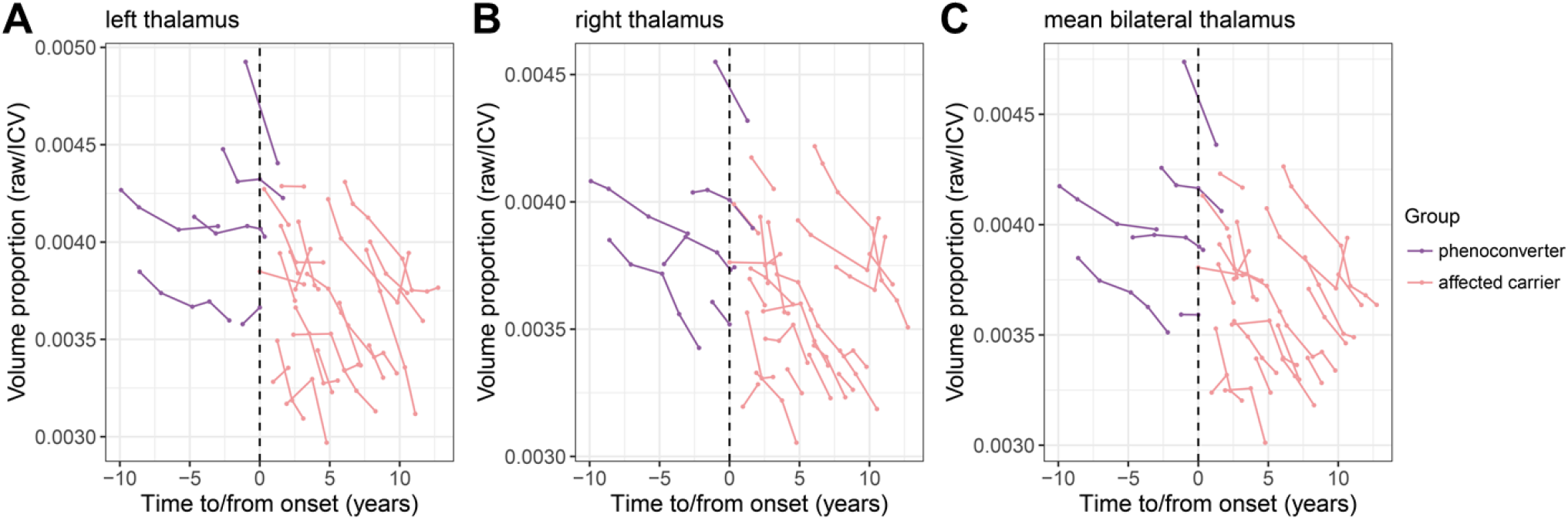
Longitudinal tracking of gray matter volume changes in thalamus. Spaghetti plot of longitudinal tracking of (**A**) left, (**B**) right, and (**C**) mean bilateral thalamic volume changes relative to time to/from symptom onset. Each scan is represented by a dot and repeated scans are connected by lines, representing the individual longitudinal changes.

## Discussion

In this study, we leveraged multi-center MRI data to conduct both cross-sectional comparisons and longitudinal tracking, aiming to examine the cortical and subcortical structural changes in persons with a *C9orf72* repeat expansion. Our findings provide evidence that brain structural changes are detectable when individuals are pre-symptomatic and later phenoconverrt to clinically manifest ALS or FTD. Smaller gray matter volume, particularly in the thalamus, was consistently observed in pre-symptomatic carriers, phenoconverters, and affected carriers, with phenoconverters demonstrating an intermediate level. Of critical importance is the observation that thalamic volume among phenoconverters was lower while still pre-symptomatic compared to those who have not yet phenoconverted; and that thalamic volume reliably predicts those with a *C9orf72* expansion who do and do not progress to phenoconversion. Overall, these findings underscore the potential utility of neuroimaging biomarkers, especially thalamic volume, as a susceptibility/risk biomarker predicting phenoconversion to clinically manifest disease, either ALS or FTD.

The pathogenic hexanucleotide repeat expansion in the *C9orf72* gene is the most common genetic cause of ALS and/or FTD, with those found to have the expansion via predictive testing being uncertain whether they will develop ALS, FTD, or a combination of both. Previous studies have highlighted differences between those with a *C9orf72* repeat expansion relative to apparently sporadic ALS-FTD spectrum disease^19^. For example, affected individuals with a *C9orf72* repeat expansion tend to exhibit more widespread cortical thinning and thalamic volume decline^20^, particularly the thalamic subregions connected to motor and sensory cortical areas^21^. For pre-symptomatic carriers with a *C9orf72* repeat expansion, reduced gray matter volume was already detectable at a relatively early age in cortical regions such as temporal, parietal, and frontal cortices, precentral gyrus, and cingulate, as well as in subcortical regions such as thalamus and striatum compared to non-carrier controls^22-25^. Our findings also converge with prior reports that consistently highlight early thalamic volume decline, and reinforce the vulnerability of the thalamus to *C9orf72*-related pathology. Importantly, in our multi-center study, we demonstrate that reduced thalamic volume is observable independent of whether initial phenoconversion is to clinically manifest ALS or FTD.

The thalamus stands out as a particularly sensitive region to *C9orf72*-related pathology and is highly interconnected with projections to both frontotemporal and motor cortical regions. Its vulnerability is likely due to several factors, including the accumulation of dipeptide repeat proteins (DPRs) within this region^26^. Furthermore, *C9orf72* methylation, which is highly correlated with severity of pathological burden^27^, has also been found to correlate with MRI volume of the thalamus^28^. This provides convergent neuropathological and neuroimaging evidence for the role of the thalamus in *C9orf72*-related disease. Specific thalamic subregions, such as the pulvinar, midline, mediodorsal, and lateral geniculate nuclei, have been detected with higher degree of atrophy in *C9orf72*-related disease^8,29^, with pulvinar atrophy appearing to be unique to *C9orf72*-related pathology^8,25^. Moreover, rodent models of *C9orf72* have demonstrated reduced thalamic volume in embryonic stages, further suggesting a specific vulnerability of this region^30^. Together, these findings underscore the importance of the thalamus not only as a region of interest in understanding the mechanisms of *C9orf72*-related neurodegeneration, but also as a candidate for neuroimaging biomarkers.

Recent clinical trials for *C9orf72*-associated ALS/FTD have included antisense oligonucleotides (ASOs) targeting *C9orf72* RNA transcripts, and therapies targeting downstream pathways of autophagy or neuroinflammation such as the PIKFYVE inhibitor and the LINE-1 retrotransposon inhibitor TPN-101^31^. However, most clinical trials are oriented towards the clinically manifest stage of disease, by which point significant neurodegeneration has already occurred. Their effectiveness may depend on timely intervention during the pre-symptomatic stage, underscoring the need for reliable susceptibility/risk biomarkers to predict the timing of phenoconversion to clinically manifest disease, and to guide optimal timing for therapeutic intervention^3^. NfL, an indicator of axonal degeneration, is a well-established example of a susceptibility/risk biomarker, at least among a subset of persons with a *SOD1* pathogenic variant. Among those with highly penetrant *SOD1* variants associated with rapid disease progression, elevated levels of NfL can be detected at least as far back as 12 months prior to clinical phenoconversion^32^. Consistent with prior neuroimaging work suggesting that the thalamus may serve as a more consistent and robust neuroimaging biomarker than cortical volume of *C9orf72* carrier ^33,34^, our cross-sectional analyses identified a pattern consistent with thalamic reductions across the disease continuum, from pre-symptomatic carriers to phenoconverters, and further to affected individuals at the clinically manifest stage. This cross-sectional pattern was further reinforced through our longitudinal data, in which we see a consistent slope in decline of thalamic volumes prior to phenoconversion.

On average, significant thalamic volume reduction was observed several years prior to symptom onset, with levels markedly lower than those of pre-symptomatic carriers and within the range of affected persons. This suggested that thalamic volume provides complementary information by offering earlier insights into the neurodegenerative trajectory. By integrating thalamic volume with other biomarkers such as NfL, future studies could develop multi-modal approaches to improve the accuracy of phenoconversion prediction and better define intervention timing points. For example, recent disease-age models of *C9orf72* identified that, in addition to thalamus having the largest MRI effect size, the combination of neuroimaging features with NfL and CDR Sum of Boxes achieved greater sensitivity for cross-sectionally based modelling of disease progression for FTD^35^, though manifest ALS was not considered. Notably, our findings regarding thalamic volume were at an individual-level, rather than mere differences in group averages. Beyond its role as a susceptibility/risk biomarker for prevention trials, individual thalamic volumes may, once validated, have additional utility in counseling about disease risk and prognosis. Importantly, the reproducibility of this biomarker across multiple centers highlights its potential as a reliable and consistent indicator of neurodegeneration in a multi-center study setting.

Despite its strengths, this study has several limitations. First, the relatively small sample size of phenoconverters (n = 8) limits the generalizability of conclusions regarding this critical transitional phase. Moreover, our analysis focused primarily on gray matter volume, while other MRI modalities, such as diffusion tensor imaging (DTI) or functional MRI (fMRI), could provide complementary insights into microstructural and functional changes. Future research integrating multi-modal imaging approaches may enhance the understanding of *C9orf72*-associated disease and strengthen biomarker development. Furthermore, while this study reinforces the importance of the thalamus in individuals with a *C9orf72* repeat expansion, future research should explore the subregions of the thalamus and their distinct roles in disease progression. Investigating thalamic projections to various cortical regions could provide deeper insights into the connectivity changes underlying clinical symptoms and disease progression.

In conclusion, the development and progression of ALS and/or FTD in individuals with a *C9orf72* repeat expansion is associated with regionally-specific brain volume loss. Structural changes are evident even at the pre-symptomatic stage, particularly in biologically plausible regions affected by *C9orf72* such as the thalamus. Compared to cortical regions, the thalamus demonstrates a stronger and more consistent signal, supported by its relevance to *C9orf72*-related pathobiology and the technical advantages of automated segmentation, which enhance signal-to-noise ratios. Thalamic volume declines longitudinally in those with a *C9orf72* repeat expansion, both before and after phenoconversion, encompassing transitions to ALS and/or FTD. This consistent trajectory of volume loss, combined with its robust performance across multi-center settings, position thalamic volume as a promising neuroimaging biomarker for early detection and phenoconversion risk prediction in pre-symptomatic carriers, highlighting its potential to guide timely and targeted therapeutic strategies as well as counseling strategies for *C9orf72* carriers at risk for ALS and/or FTD.

## Data availability

All datasets used and/or analyzed during the current study are available from the corresponding author on reasonable request and approval from the Penn Neurodegenerative Data Sharing Committee. Requests may be submitted using a webform request: https://www.pennbindlab.com/data-sharing.

## Acknowledgements

We extend our gratitude to patients, study participants, and their families for their invaluable contributions, which made this study possible. We also thank the Pre-fALS study team at the University of Miami for study coordination; Prof. Peter M. Andersen (Umea University) for Pre-fALS C9orf72 testing; and Christine Stanislaw (Emory University) for Pre-fALS genetic counseling. The study was supported by NIH (P01-AG06697, P30-AG072979, U54-NS092091, R01-NS105479, RF1-NS145263).

## Supplementary Figures

**Supplementary Figure 1.**
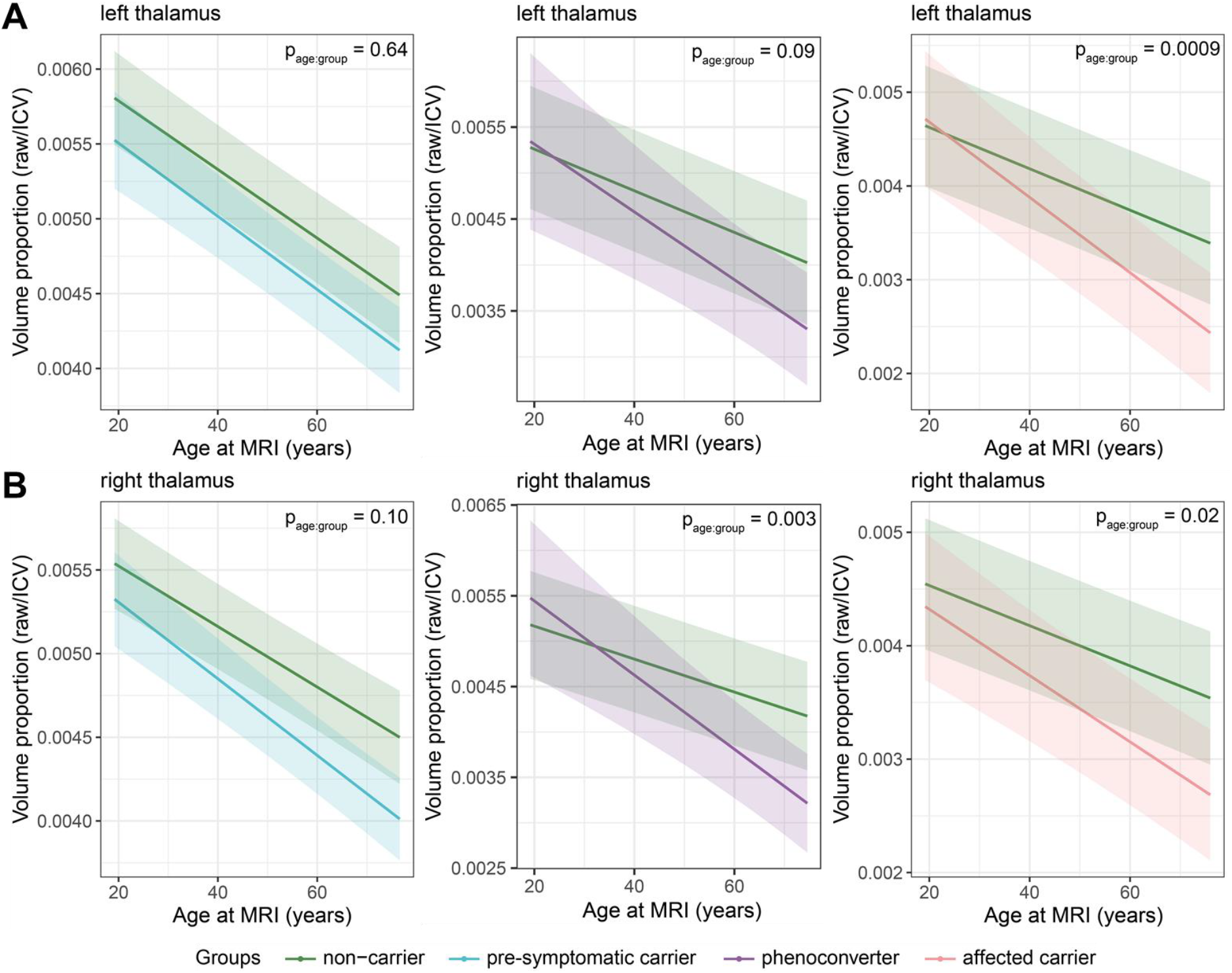
Age-related trajectories of thalamic volume by groups. Age-by-group interactions on left (A) or right (B) thalamus in different groups. Lines and shaded 95% confidence intervals from a linear mixed model accounting for sex and MRI sequence. P_age:group_ represents the significance of age-by-group interactions.

**Supplementary Figure 2.**
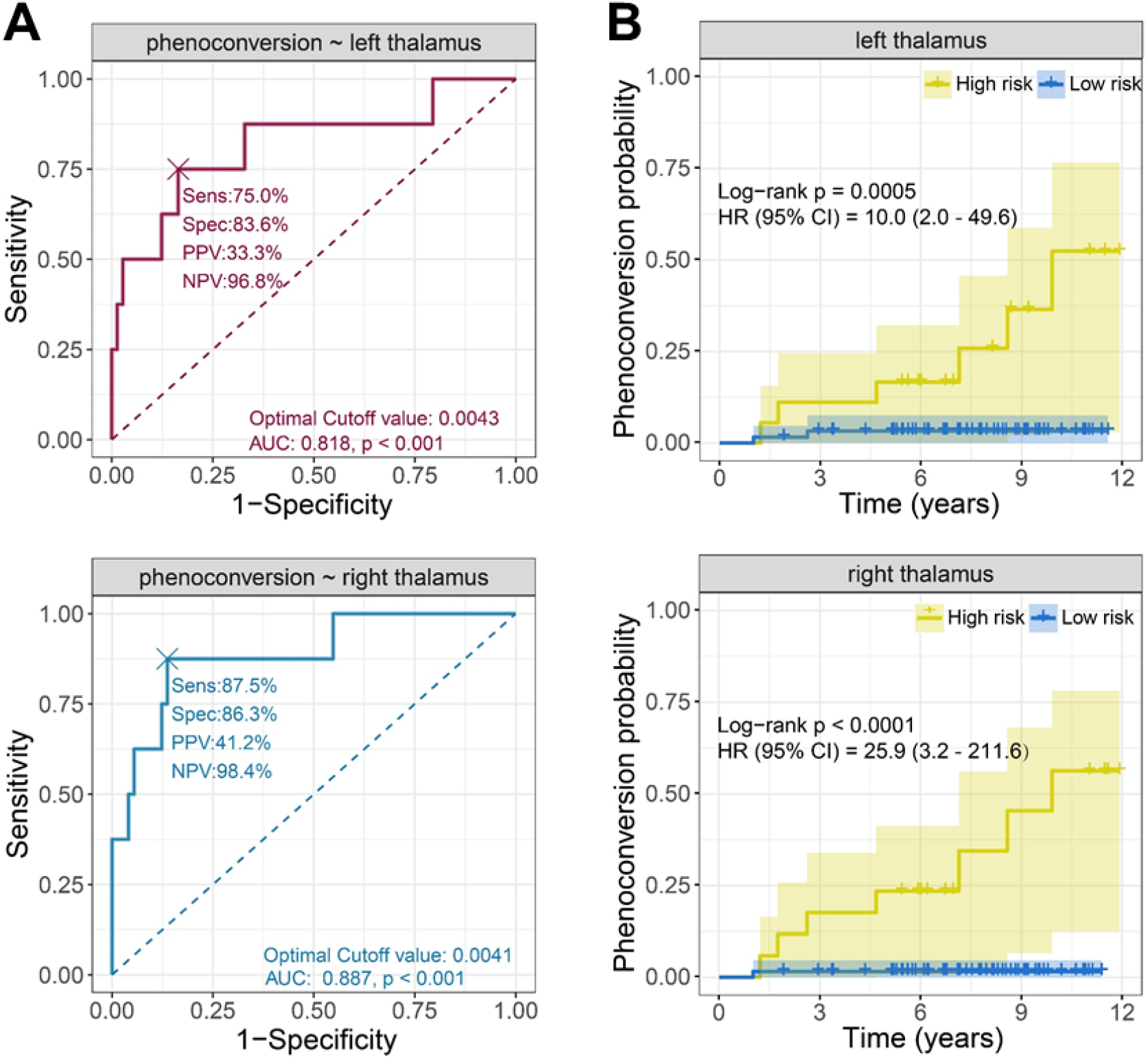
Analysis of phenoconversion prediction. (**A**). ROC curves for predicting phenoconversion using left or right thalamic volumes. The “×” mark represents the optimal cutoff point.(**B**) Kaplan-Meier curves showing phenoconversion probability by risk groups. Shading represents 95% CI. HR, hazard ratio; CI, confidence interval.

**Supplementary Figure 3.**
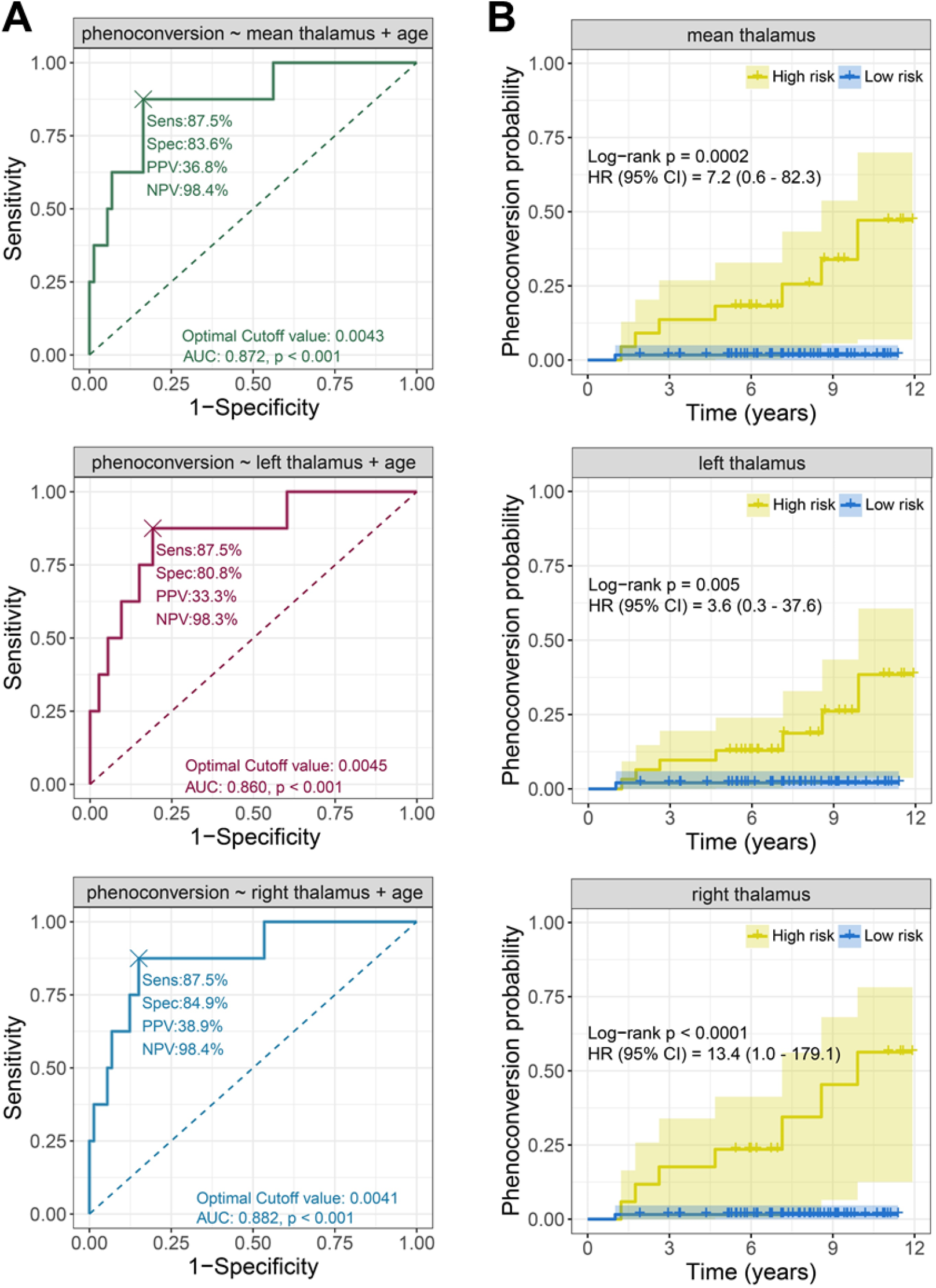
Analysis of phenoconversion prediction adjusting for age. (**A**). ROC curves for predicting phenoconversion using thalamic volumes. The “×” mark represents the optimal cutoff point. (**B**) Kaplan-Meier curves showing phenoconversion probability by risk groups. Shading represents 95% CI. HR, hazard ratio; CI, confidence interval.

